# Forecasting American COVID-19 Cases and Deaths through Machine Learning

**DOI:** 10.1101/2020.08.13.20174631

**Authors:** Anaiy Somalwar

## Abstract

COVID-19 has become a great national security problem for the United States and many other countries, where public policy and healthcare decisions are based on the several models for the prediction of the future deaths and cases of COVID-19. While the most commonly used models for COVID-19 include epidemiological models and Gaussian curve-fitting models, recent literature has indicated that these models could be improved by incorporating machine learning. However, within this research on potential machine learning models for COVID-19 forecasting, there has been a large emphasis on providing an array of different types of machine learning models rather than optimizing a single one. In this research, we suggest and optimize a linear machine learning model with a gradient-based optimizer for the prediction of future COVID-19 cases and deaths in the United States. We also suggest that a hybrid of a machine learning model for shorter range predictions and a Gaussian curve-fitting model or an epidemiological model for longer range predictions could greatly increase the accuracy of COVID-19 forecasting.

## 1 Introduction

COVID-19, the disease caused by the virus SARS-CoV-2, in the past six months, has been the cause of death for over 130,000 in the United States [1]. On February 3rd, 2020, the United States government declared COVID-19 a Public Health Emergency, just three days after the World Health Organization (WHO) declared COVID-19 a Global Health Emergency [2]. A little over one month later, on March 6th, 2020, COVID-19 was declared a National Emergency in the United States, which allowed for the additional funding for the research on COVID-19 [2]. With both the United States and international total COVID-19 cases and deaths increasing at a rapid rate, models that predict the growth and decline of the coronavirus are ever so important.

In the modelling of most epidemics and outbreaks, the accuracy of the predictions of the future number of cases and deaths has largely depended on the amount of relevant, accessible data. This amount of relevant data can be measured in two metrics - the length of the data in days, and the width of the data in relevant categories. The traditional predictive modelling of epidemics and outbreaks often face the same challenges; the small “length” of data from the beginning, and a width of data that contains several irrelevant categories. Even as the duration of the outbreak continues, there may be some inputs that are especially relevant to the predictions for the future cases and deaths of the outbreak that are not being tracked or available to the public.

Most traditional epidemiological models contain one common characteristic, a system of equations that is used to replicate the human to human transmission of a disease [4, 8]. However, after March 27th, 2020, when the Institute of Health Metrics and Evaluation (IHME) at the University of Washington described their method of predictions for COVID-19 deaths and healthcare demand per state in a preprint on medRxiv [3], there has been much more attention on a different type of COVID-19 forecasting model, a model that fits a postulated Gaussian distribution of COVID-19 deaths. While these types of models are currently being developed and improved upon, there has been much less focus on simple, autoregressive models for COVID-19 forecasting even within machine learning based models. The goal of our research is to examine how a primarily autoregressive machine learning model will compare with traditional epidemiological models and the current state of the art models and whether machine learning models could be the optimal solution for COVID-19 forecasting.

## 2 Methods

In this paper, we focus on predicting future COVID-19 cases and deaths in the United States as those metrics seem to be the most relevant based on the previous research on COVID-19 forecasting and their general applications in healthcare and policy making. We chose to use the national data for the United States from the New York Times’ Github under the covid-19-data repository [6] as it was updated daily and contained no missing values. This dataset contains three columns with the date, total number of cases of COVID-19 in the United States, and the total number of deaths due to COVID-19 in the United States since the first confirmed COVID-19 case in the United States on January 21, 2020.

To predict the future total United States COVID-19 cases or deaths, we assume that there exists functions *f* and *g* that respectively map the COVID-19 cases and deaths of some past constant *i* days to the COVID-19 cases and deaths of the next day, or the *i* + 1th day. This can be seen in the equations below, where *c_k_* represents the number of cases at index *k, d_k_* represents the number of deaths at index *k, t* represents the index of the current day, and *n* and *k* are the respective number of previous days inputs for the prediction of future cases and deaths.

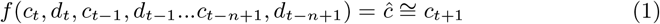

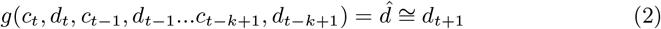

To create these models, we downloaded and pre-processed the data from the New York Times’ github using several common Python libraries to create the input and output arrays described above. We split the first 80 percent of the chronologically sorted data to serve as training data where the model “learns” the function that mapped the inputs to the outputs, and we used the remaining 20 percent of my data to test how well the model would perform for data it had never seen. We found that a simple linear regression model with two cross-validation sets and 12 penalization (Ridge Regression) was performing the best after we set *n* = 7 and *k* = 5. The models can be described by the equation below, where *b*_0_ and 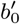 are biases and *w*_1_ to *w*_2_*_n_* as well as 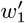 to 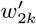 are individual weights.

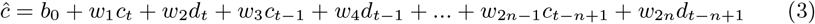

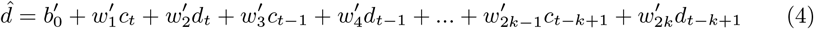

The model fits on the training data by minimizing a loss function, which we have chosen to be mean squared error plus an l2 regularization term, which is intended to guide the model to having relatively small weights to prevent overfitting. The exact loss function can be seen below, where 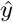 is an array of predictions with size *n*, *y* is an array of real values with size *n*, λ is a constant, and *θ* is an array of weights.

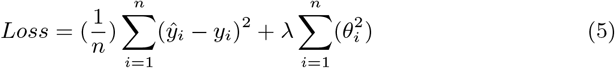

To predict the next *l* future cases or deaths from current time *t*, we recursively use our models’ output of the cases and deaths for time *t* + 1 to serve as input for predicting the cases or deaths for time *t* + 2 and so forth. We chose *l* as 7 while making predictions as that was a number which was used in previous literature on this topic and made this model easily comparable. We made six *l* day predictions of for every *l* day interval of the testing data, which started on 6/27/20.

## 3 Results

We measure the model’s error by the square root of the mean squared error (RMSE) of its predictions on the testing data, which represents the average number of cases or deaths the prediction is off by.

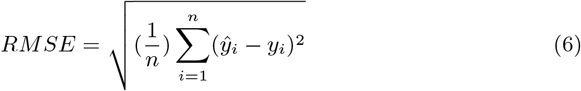

We also measure the model’s accuracy based on its *r*^2^, or its coefficient of determination, which is a statistical measure of how close a regression is to real values.

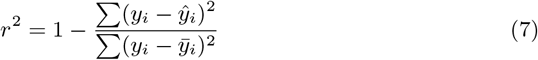

The Ridge Regression recursive model for predicting future United States COVID-19 total deaths performs with an average RMSE of 570.264 based on the average RMSE of its seven-day prediction task. The Ridge Regression recursive model for predicting future United States total COVID-19 cases performs with an average RMSE of 4282.341 based on the average RMSE of its prediction task. The *r*^2^ value for the recursive deaths predictor is 0.996, and the *r*^2^ value for the recursive cases predictor is 0.992. Figures 1 and 2 depict the accuracy of the models’ predictions.

**Figure 1:**
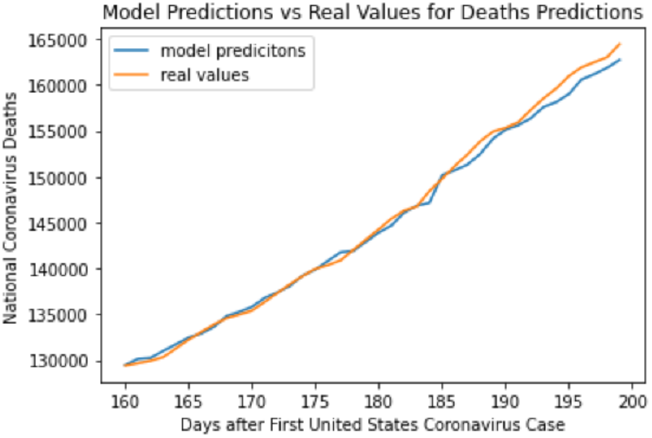
A visualization of the Ridge Regression’s predictions on the United States total of coronavirus deaths.

**Figure 2:**
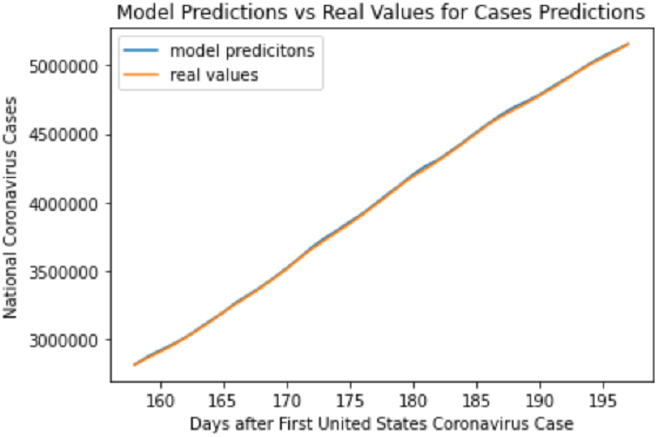
A visualization of the Ridge Regression’s predictions on the United States total of coronavirus cases.

## 4 Discussion

As can be seen the *r*^2^ values of the Ridge Regression for the predictions for future COVID-19 cases and deaths, the model fits well on the testing data for both prediction tasks. We compare this model to several ones in the literature, namely the epidemiological model from [5] and the Gaussian curve-fitting model from [7].

As can be seen in the accuracy statistics for the exponential smoothing model from [5] on the United States, the lowest RMSE for the cases predictions was a little under 6000, whereas the Ridge Regression’s average RMSE across the end of its six, seven day prediction tasks was 4282.341. While the Ridge Regression’s predictions were on a later testing data, which indicates a larger amount of training data, the total number of COVID-19 cases in the United States had greatly increased, which makes its smaller RMSE more meaningful in terms of its percent error. However, it is important to note that the prediction task of the exponential smoothing model was a 10 day interval, which is a more difficult task. Nonetheless, it is clear that the Ridge Regression model is competitive with the traditional epidemiological exponential smoothing one.

Since our has shown that it is competitive with a traditional epidemiological model, we compare it to the current state of the art Gaussian curve-fitting models for United States COVID-19 death prediction tasks such as those from the IHME and the University of Texas at Austin’s COVID-19 Modelling Consortium. As described in the University of Texas at Austin’s COVID-19 Modelling Consortium’s preprint[7], their model performed significantly better than that of the IHME, which is the model that was widely used in healthcare and policy making. The improved model averaged a 13.2 average state RMSE for 13 day predictions of total United Stats COVID-19 deaths, or a 660 average national RMSE. While the Ridge Regression model for deaths has a lower absolute RMSE on national deaths, it should be taken into account that the size of the training data was larger for the Ridge Regression and the Ridge Regression performed well on a smaller prediction interval and a more simple prediction task. However, machine learning models such as the Ridge Regression undoubtedly perform competitively on shorter prediction tasks.

## 5 Conclusion

In the United States as well as many other countries, COVID-19 has caused a national state of emergency and a great healthcare problem. While this paper improves upon the existing machine learning models for this pandemic, it is clear that there is space for some improvement on the longer-range predictions of machine learning models. However, machine learning does seem to be a competitive option for COVID-19 modelling and a hybrid of curve-fitting models and machine learning models seems promising for COVID-19 forecasting.

## Data Availability

The datasets generated during and/or analysed during the current study are available in the covid-19-data repository in the New York Times' Github.

https://github.com/nytimes/covid-19-data

